# Predicting Melanoma Staging using Targeted RNA Sequencing data using Machine Learning

**DOI:** 10.1101/2021.11.03.21265077

**Authors:** Fahad Shabbir Ahmed, Furqan Bin Irfan

**Author notes:** Corresponding author: Fahad Shabbir Ahmed, MD Clinicaro Machine Learning Group, New Haven, CT 06510, United States; Department of Pathology, Wayne State University, Detroit, MI 48201, United States. Corresponding author contact details.

## Abstract

The aim of this study is to use machine learning to predict tumor staging and metastasis in melanoma with differentially expressed genes. Machine has been used in different clinical setting to predict different outcomes. However, it has not been used to look at predicting the diagnostic aspect of tumor staging. We used the TCGA RNA-Sequencing data on melanomas to predict tumor staging nodal and/or metastasis using deep neural networks (DNN) and random forest classifier (RF). Results: We were able to predict tumor staging (lower vs higher stage, i.e. Tis / T1 / T2 vs T3 and higher), nodal metastasis and combined nodal or distant metastasis in patients with melanomas with high accuracies. However, we need to further validate these results.

## INTRODUCTION

Cancer Staging tends to be a lengthy and expensive process with an overall out of pocket cost reaching 5.6 billion USD in 2018^1^. Tumor staging is an integral part of patient care, and it directly correlates to what treatment should be used in these patients. We have previously shown that a pan-cancer machine learning algorithm can predict nodal metastasis utilizing differentially expressed genes (DEGs) from RNA-sequencing data^2^ Cancer metastasis is a late-stage condition of the disease. Machine learning has the potential to predict clinical outcomes in other diseases allowing earlier management and prevention of complications^3,4^. In our current study, we hypothesize that DEGs can be used as a biomarker to predict tumor staging, metastasis (nodal and distant) in melanomas.

The use of machine learning (ML) has been used in multiple industries including financial, defence, manufacturing and over the past decade. While the use of ML in medicine and specifically in cancer diagnostics has been limited to radiological imaging technology and has provided guidance for how approvals can take place^5^. In the recent publications we have successfully used ML to predict mortality as a clinical outcome in the in-patient and intensive care setting with good success^6,7^.

The TNM Staging of cancer is critical in making the most informed choices for clinical treatment of these patients. After the first biopsy is done and a cancer diagnosis is made based on histopathology and relevant immunocytochemistry; it takes a few weeks until a CT and or PET scans can be done for staging the tumor after getting approval from insurance and availability of slots for the scans. For these patients time is a precious commodity and here in this paper we wanted to develop a tumor staging model for patients with melanomas as a starting point. This work is based on our pan-cancer nodal metastasis work published previously using ML^8^.

## METHODS

### Patient selection criteria

The Cancer Genome Atlas (TCGA) has accumulated RNA-sequencing transcriptome data from thousands of cancer tissue samples across 33 cancer types^9^. We identified DEGs and corresponding clinicopathological characteristics for melanoma cases in the TCGA database. RNA Sequencing data for melanoma cases was extracted from the TCGA database.

### Pre-processing of data

All clinical information of the cases were aligned with the respective sequencing data. 203 genes of interest were identified and an upper and lower limit cut-offs for the sequencing data were used to classify genes as either high or low expression.

### Machine Learning Experiments

We conducted six different machine learning experiments for this paper. The first experiment was conducted to look at if pan-expression without cut-offs could predict nodal metastasis. The second and third experiment evaluated if machine learning could be used to predict nodal metastasis by looking at 203 genes with upper and lower limit cut-offs with deep neural networks (DNN) and random forest classifier (RF). The fourth model evaluated if distant site metastasis (non-nodal) could be evaluated using RF. Fifth model looked at prediction of combined metastasis (nodal and/or distant metastasis). The sixth and final model looked at prediction of lower stage (Tis, T1 and T2) vs higher stage (T3 and T4). All datasets were split using 80/20 for training and test sets. Synthetic Minority Oversampling Technique (SMOTE) was utilized to address the imbalanced distribution of the outcome in the dataset. The algorithm accuracy was determined by sensitivity, specificity, predictive values (positive and negative), and area under the receiver-operator curve (AUROC).

### Software

R-Language and Microsoft Excel 365 was used for data wrangling. Machine learning models were trained and evaluated using Python 3.8.

## RESULTS

RNA-sequencing samples from 490 melanoma patients that were analysed from the TCGA database. For prediction of tumor stage (stage III or higher), nodal metastasis, distant metastasis and combined nodal or distant metastasis were considered. The results for nodal metastasis with total gene expression data, Nodal Metastasis (Pan-Expression) Deep Neural Network versus selected gene expression data (Targeted Expression) Deep Neural Network showed sensitivity of 66.22% vs 81.13%; specificity of 16.98% vs 100.0; positive predictive value 52.69 vs 100.0; negative predictive values of 26.47 vs 88.10; training set accuracies of 57.29 vs 100.0; test set accuracies of 45.67 vs 92.13; and AUROCs (95%CI) of 0.495(0.25-0.47) vs 0.982 (0.62-0.83). Another algorithm for nodal metastasis; (Targeted Expression) Random Forest, had outperformed the previous one and showed sensitivity (97.89), specificity (100.00), PPV (100.00), NPV (94.00) and AUROC (1.00, 95%CI 0.91-0.99). The use of DEG for the distant metastasis staging algorithm; Distant Metastasis (Targeted Expression) RF, showed sensitivity (0.00), specificity (100.00), PPV (0.00), NPV (100.00) and AUROC (1.00, 95%CI 0.91-1.00). While DEG for combined metastasis (nodal or distant) staging algorithm; Nodal or Distant Metastasis (Targeted Expression) Random Forest, showed sensitivity (97.56), specificity (100.00), PPV (100.00), NPV (98.56) and AUROC (1.00, 95%CI 0.88-0.98). The tumor staging (DEG and predicting higher stage i.e stage 3 or higher) algorithm; Tumor Staging (Targeted Expression) Random Forest, showed sensitivity (100.00), specificity (100.00), PPV (100.00), NPV (100.00) and AUROC (1.00, 95%CI 0.69-0.89).

## DISCUSSION

Our work proves for the first time that machine learning-based differential gene expression for cancer metastasis prediction is possible. The targeted machine learning models for differential gene expression show better performance for metastasis prediction in melanoma than the complete pan-gene expression model. Moreover, machine learning can predict nodal and distant metastasis with high accuracy and can be combined in a single algorithm that can predict either nodal or distant metastasis. These algorithms can directly be used to determine staging of the cancer, if the machine learning targeted gene expression shows metastasis in melanoma, then we can consider stage 3 or 4 (Table 1). The staging can be confirmed with imaging modalities as is currently the case. The clinical utility of such a diagnostic approach using machine learning-based differential gene expression will provide immediate results for metastasis at the same time of histopathology proven cancer diagnosis. The patient and the physician will know the metastasis status at the same time as diagnosis, management can be planned and started much early. And these results can be confirmed with imaging.

**Table 1.** Machine learning models

| Experiment | Sensitivity | Specificity | PPV | NPV | AUROC | 95%CI | Test Set Accuracy | N |
| --- | --- | --- | --- | --- | --- | --- | --- | --- |
| Nodal Metastasis (Pan-Expression) DNN | 66.22 | 16.98 | 52.69 | 26.47 | 0.495 | 0.25-0.47 | 54.55 | 479 |
| Nodal Metastasis (Targeted Expression) DNN | 81.13 | 100.0 | 100.0 | 88.10 | 0.982 | 0.62-0.83 | 90.21 | 479 |
| Nodal Metastasis (Targeted Expression) RF | 97.89 | 100.00 | 100.0<br>0 | 94 | 1.00 | 0.91-0.99 | 96.74 | 458 |
| Distant Metastasis (Targeted Expression) RF | 0.00 | 100.00 | 0.00 | 100 | 1.00 | 0.91-1.00 | 94.44 | 450* |
| Nodal or Distant Metastasis (Targeted Expression) RF | 97.56 | 100.00 | 100.0<br>0 | 98.56 | 1.00 | 0.88-0.98 | 98.91 | 458 |
| Tumor Staging (Targeted Expression) RF | 100.00 | 100.00 | 100.0<br>0 | 100.0<br>0 | 1.00 | 0.69-0.89 | 100.00 | 395 |

\*Only 27 cases of distant metastasis were identified.

These results need further external validation. We also need bigger sample sizes for creation and confirmation of machine learning-based differential gene expression for metastasis prediction. For example, only 27 cases of distant metastasis for melanoma were available in the TCGA database. Future direction also includes prospective trials to confirm the results. Development of such machine learning-based differential gene expression to predict metastasis is currently underway for breast cancer, genitourinary (prostate, ovarian, etc.), gastrointestinal (colon, pancreases, etc.) respiratory tract (lung, head and neck, etc.), Neuronal (central and peripheral nervous system) and haematological (lymphomas and leukaemia) cancers.

Machine learning-based differential gene expression for cancer metastasis prediction has the potential to change the approach and management of cancer patients. Such a machine learning-based differential gene expression diagnostic assay for metastasis can save valuable time for cancer patients and start treatment weeks and even months before the imaging results are available for metastasis.

## CONCLUSION

Our machine learning models can predict tumor staging including higher vs lower stage tumor, nodal metastasis and combined metastasis with high accuracy. However, these results need to be further validated with a new prospective independent external validation set to be ready as a clinical tool.

## Data Availability

available on request

---

**Figure 1.**
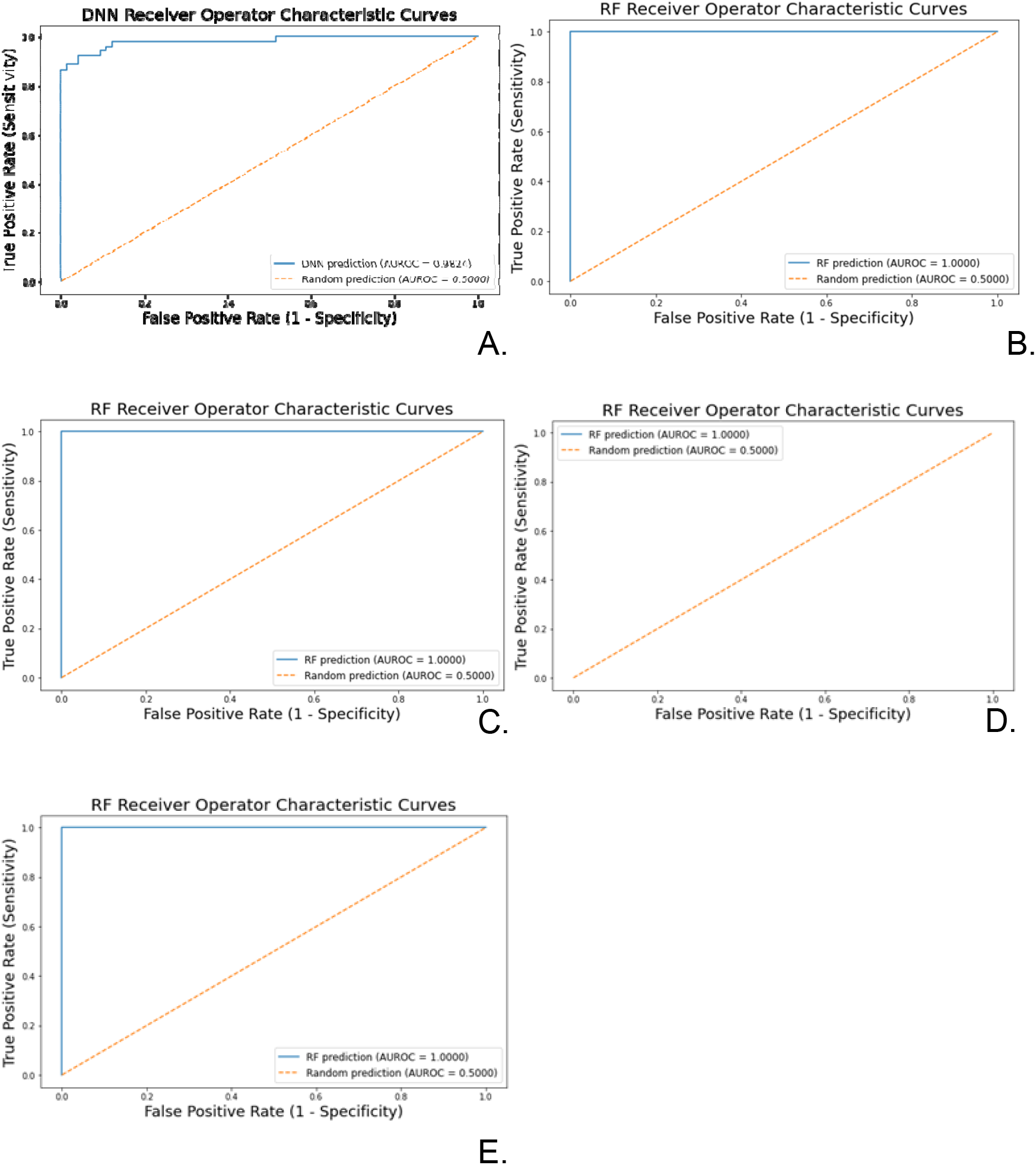
A. Targeted expression nodal metastasis prediction using DNN. B. Targeted expression nodal metastasis prediction using RF. C. Targeted expression distant metastasis prediction using RF. D. Targeted expression nodal or distant metastasis prediction using RF. E. Targeted expression for tumor staging prediction (Stage 3 or higher).

